# Serum Metabolomic Changes Following Bariatric Surgery Adjusted for Measured Glomerular Filtration Rate Suggest Mechanisms of Kidney Protection

**DOI:** 10.1101/2025.05.30.25328578

**Authors:** Benjamin Apple, Jingsha Chen, Tooraj Mirshahi, Christopher Still, Andrew S. Levey, Lesley Inker, Morgan Grams, Josef Coresh, Evan M Zeitler, Alexander R Chang

## Abstract

**Background:** Bariatric surgery reduces glomerular hyperfiltration in the short term and is associated with a reduced risk of glomerular filtration rate decline during long-term follow-up. Assessing surgery-induced changes in serum metabolites may be useful to understand the metabolic benefits to the kidney occurring after bariatric surgery.

**Methods:** In a prospective, single-center research cohort of 27 adults with severe obesity who underwent bariatric surgery, we measured serum metabolites using untargeted ultrahigh performance liquid chromatography-tandem mass spectrometry and measured glomerular filtration rate (mGFR) by iohexol plasma clearance 1-3 months prior and 6 months after bariatric surgery. In generalized estimating equation (GEE) models that included age, sex, mGFR, and post-surgery terms, we examined bariatric surgery-associated changes in serum metabolites as well as associations between serum metabolites and mGFR. We used MetaboAnalyst to perform pathway analyses to determine bariatric surgery-associated metabolite pathway changes.

**Results:** Bariatric surgery was significantly associated with changes in 223 serum metabolites after adjustment for age, sex, and mGFR at a Bonferroni-corrected p-value of 4.85 × 10^−5^. Following bariatric surgery, there were several pathways that were downregulated (alpha-linoleic acid and linoleic acid, methionine, tyrosine-kynurenine, and alanine-glucose metabolism pathways; raw p<0.05) or upregulated (phenylacetate, bile acid biosynthesis, taurine and hypo-taurine metabolism, porphyrin metabolism pathways; raw p<0.05), though only downregulation of alpha-linoleic acid and linoleic acid metabolism pathway was significant after correcting for multiple comparisons. Creatinine also demonstrated a significant mGFR-independent decrease following surgery. Top metabolites significantly associated with mGFR included N,N,N-trimethyl-alanyl proline betaine (TMAP), followed by creatinine, N-acetylethreonine, pseudouridine, N-acetylserine, myo-inositol, 5-methylthioribose, 5,6-dihydrouridine, erythronate, and N6-succinyladenosine.

**Conclusion:** We confirmed several mGFR-independent metabolomic changes after bariatric surgery and also identified metabolites associated with mGFR in this setting. Further studies are needed to investigate the potential mechanistic role of identified metabolites to clarify mechanisms of obesity-related kidney disease.

## INTRODUCTION

Bariatric surgery is one of the most effective weight loss interventions for individuals living with obesity.(1, 2) In addition to weight loss, bariatric surgery is associated with a multitude of salutary metabolic changes.(1, 2) Following bariatric surgery, chronic metabolic diseases, including type II diabetes mellitus, hypertension, dyslipidemia, and fatty liver disease, may go into long-term remission.(1–3)

Metabolite changes following bariatric surgery correlate with the observed clinical improvements in disease.(1, 3) For example, serum metabolites associated with insulin resistance significantly decrease following surgery, whereas, metabolites associated with insulin sensitivity significantly increase following surgery.(1, 3–5) Additionally, metabolites associated with inflammation, such as sphingomyelins, long-chain fatty acids, and ceramides significantly decrease following surgery, suggesting a decrease in chronic inflammation.(1, 3, 4) However, the available studies investigating metabolomic changes following bariatric surgery fail to adjust for changes in kidney function following surgery, a critical limitation given that GFR is associated with a wide variety of measured metabolites (6).

Reductions in GFR are associated with the accumulation of a host of metabolites compared to normal kidney function. Individuals with chronic kidney disease (CKD) demonstrate increased levels of inflammatory metabolites of tryptophan metabolism, including kynurenine pathway metabolites and indoxyl sulfate, compared to individuals with healthy kidney function.(7–10) Patients with end stage kidney disease (ESKD) demonstrate increases in uremic toxins, such as p-cresol sulfate and trimethylamine N-oxide (TMAO), compared to individuals with normal kidney function.(11, 12)

Bariatric surgery is associated with a long-term improvement in kidney outcomes. (13, 14) However, changes in creatinine-based estimated GFR (eGFRcr) after bariatric surgery are difficult to interpret because of confounding by changes in lean mass.

Studies using measured GFR (mGFR) and eGFR based on filtration markers less affected by muscle reveal consistent findings.(14) Despite a short-term decrease in glomerular filtration rate (GFR), individuals with obesity who undergo bariatric surgery have a slower rate of GFR decline and lower incidence of end-stage kidney disease (ESKD) up to 9 to 10 years post-surgery when compared to individuals with obesity who do not undergo surgery.(13) While the mechanism of benefit remains unclear, multiple lines of evidence suggest that alterations in metabolic mediators may play a role.

Few to no studies have investigated the interplay between the metabolomic changes that follow bariatric surgery and the associated improvement in kidney function outcomes. To better understand the mechanisms by which bariatric surgery-induced metabolic changes impact kidney outcomes after bariatric surgery, we aimed to identify changes in serum metabolites following bariatric surgery independent of mGFR.

## METHODS

### Study Population

The study population included 27 participants ≥ 18 years with a BMI ≥35 kg/m^2^ who were recruited in the Geisinger Center for Nutrition and Weight Management during bariatric surgery evaluation, consented for this research study and completed a pre-surgery and 6-month follow-up mGFR study visit (15, 16). Exclusion criteria included allergy to iodine or contrast dye, pregnancy, eGFRcr < 30 mL/min/1.73m2), end-stage kidney disease (ESKD), history of kidney transplant, autoimmune thyroiditis, cirrhosis, and active cancer treatment. The study was approved by the Geisinger Institutional Review Board (IRB Number 2014-0293). All participants provided written informed consent.

### Metabolomic Data Acquisition

Fasting serum samples stored at −80°C following collection at 1-3 months before and at 6 months after bariatric surgery. Untargeted metabolomic profiling was conducted by Metabolon (Morrisville, NC) using GC-MS and ultra-high-performance liquid chromatography-tandem MS on a Thermo Scientific Orbitrap MS analyzer as previously described. (17) Metabolites were categorized into 9 superpathways: amino acids, lipids, carbohydrates, TCA cycle, nucleotides, cofactors and vitamins, peptides, partially characterized molecules, and xenobiotics. Of these superpathways, metabolites were further divided into 118 subpathways.

### Measured Glomerular Filtration Rate (mGFR) Calculation

GFR was measured 1-3 months prior to surgery and 6 months after surgery as follows: 5 mL of iohexol was administered intravenously over 30 seconds, followed by 10 mL of normal saline. Blood samples were drawn at 10, 30, 240, and 300 minutes in a fashion similar to that used in the Multi-Ethnic Study of Atherosclerosis (MESA) Kidney study. (18) GFR was calculated using plasma iohexol clearance, using all time points in a 2-compartment model. Because body surface area (BSA) may change after bariatric surgery, GFR was reported without indexing for BSA.

### Statistical and Pathway Analysis

Generalized estimating equations (GEEs) were used to assess associations between serum metabolite levels and bariatric surgery and to determine associations between serum metabolite levels and mGFR. Metabolite data was normalized using log2 transformation. GEE models included age, sex, mGFR, and a dummy variable for post-surgery visit. Sensitivity analyses additionally adjusted for diabetes and BMI. Metabolites were grouped by superpathways and subpathways, and metabolites were deemed to have a significant change post-surgery or significant association with mGFR at Bonferroni-adjusted p-value (p<4.85E-5) or FDR p-value < 0.05. Generalized Estimating Equation analyses were conducted using Stata software version 15.1.

Metaboanalyst 5.0 was used to conduct pathway analysis using metabolite set enrichment analysis (MSEA) to identify metabolic pathway changes pre-to-post bariatric surgery. Each metabolite and the associated fold-change and p-value from pre-to-post bariatric surgery was submitted to the pathway analysis tool. Over Representation Analysis (ORA) was used to determine whether given metabolite sets were over-represented in the upward or downward direction from pre-to post-surgery. One-tailed p-value of 0.05 was used to determine significant over representation of metabolic pathway change in upward direction or downward direction following surgery. Holm test and FDR p-value were also used to account for multiple comparisons.

## RESULTS

### Study population demographics

Of the 27 study participants, 20 underwent Roux-en-Y gastric bypass (74.1%), 4 underwent biliopancreatic diversion with duodenal switch (BPD-DS) (14.8%), and 3 underwent gastric sleeve (11.1%). Mean age was 46.7 years, 18 (66.7%) were female (66.7%), and most were white (96.3%). At 6 months following surgery, the study group had a median BMI decrease of 13.9 kg/m^2^ and body surface area (BSA) decrease of 0.30 m^2^ (**Table 1**). 8 of 11 patients with type 2 diabetes at baseline demonstrated diabetes remission (defined by normalization of hemoglobin A1c) and 6 of 10 patients with hypertension demonstrated normalization of blood pressure (defined as systolic blood pressure <140 mmHg and diastolic blood pressure <90 mmHg without antihypertensive medication use) (**Table 1**). Additionally, the group’s median mGFR decreased from 117.3 mL/min (SD 34.1) before surgery to 108.2 mL/min (SD 24.2).

**Table 1.** Pre-surgery and post-surgery characteristics of study population. Continuous variables are reported as mean (standard deviation) and categorical variables are reported as number (percent). BSA was calculated using the DuBois equation.

|  | **Pre-surgery** | **6 months post-surgery** |
| --- | --- | --- |
| **Age** | 46.2 (10.8) | 47.1 (10.8) |
| **Female Sex** | 18 (66.7%) | 18 (66.7%) |
| **Black Race** | 1 (3.7%) | 1 (3.7%) |
| **Type II Diabetes Mellitus** | 11 (40.7%) | 3 (11.1%) |
| **Hypertension** | 16 (59.3%) | 10 (37.0%) |
| **BMI (kg/m<sup>2</sup>)** | 49.5 (9.4) | 35.6 (6.6) |
| **BSA (m<sup>2</sup>)** | 2.42 (0.27) | 2.12 (0.25) |
| **mGFR (ml/min)** | 117.3 (34.1) | 108.2 (24.2) |

### Serum metabolites associated with mGFR

Of all serum metabolites measured before and after surgery, 27 demonstrated a significant association with mGFR when using the Bonferroni-adjusted p-value (4.85E-5) and FDR p-value (0.05). Most metabolites were negatively associated with mGFR. The serum metabolites most significantly associated with mGFR were products of amino acids metabolism (4 of the top 10). N, N, N-trimethyl-alanylproline betaine (TMAP) (beta: −0.734, p-value: 2.01E-12) demonstrated the most significant association with mGFR out of all the metabolites studied (**Table 2**). Creatinine (beta: −0.341, p-value: 7.54E-10) was the next most significant metabolite associated with mGFR. When using Bonferroni-adjusted p-values (4.85E-5), eight serum metabolites demonstrated a significant change post-surgery and were significantly associated with mGFR (**Table 2)**. In addition to being strongly inversely associated with mGFR level, TMAP (beta for mGFR: −0.733, p-value 2.01E-12) significantly decreased following bariatric surgery (beta for surgery: −0.350, p-value 3.57E-8). Creatinine (beta: −0.199, p-value 1.92E-23) also demonstrated a significant decrease following bariatric surgery.

**Table 2.**
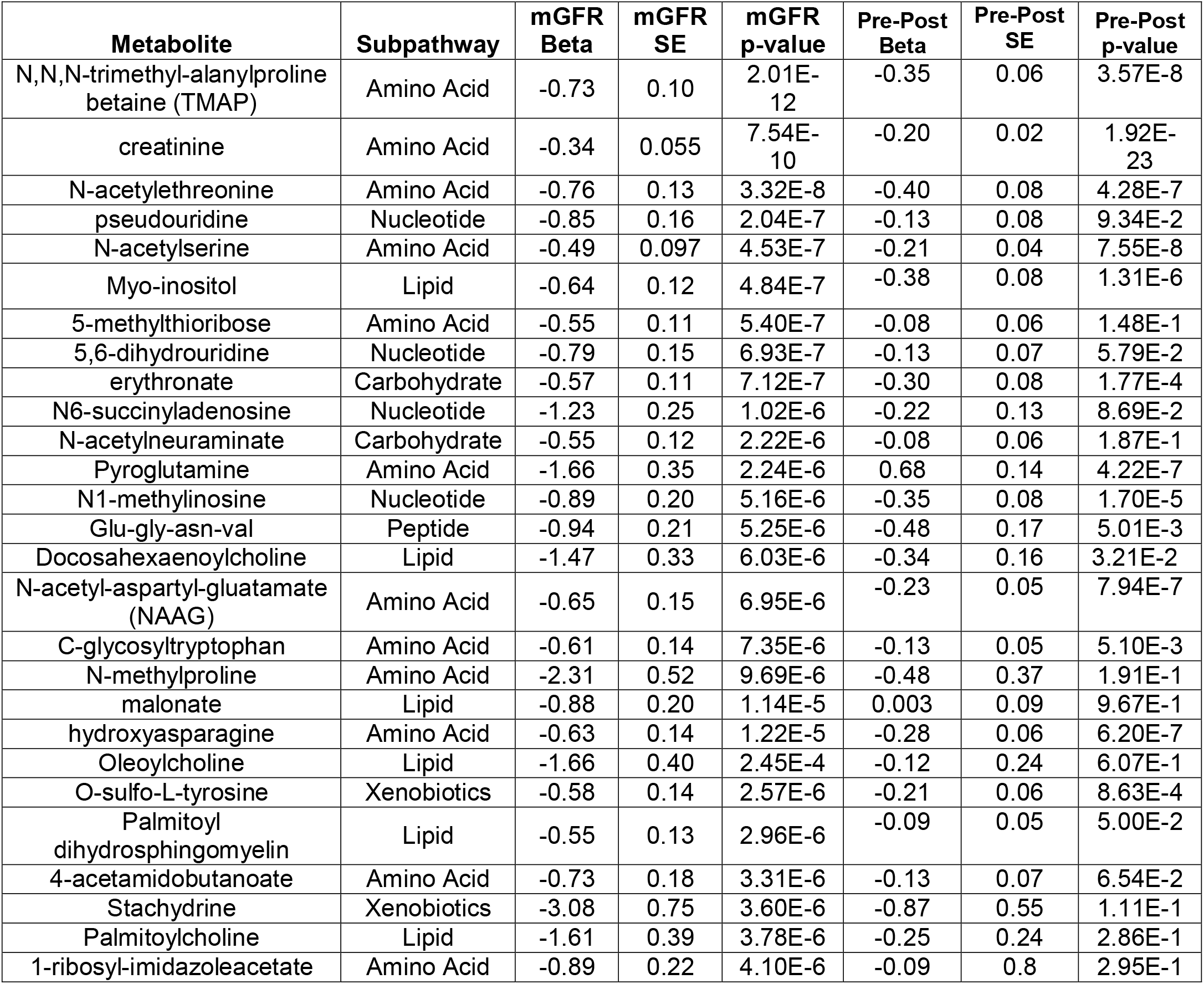
Serum metabolites significantly associated with mGFR when adjusted for age, sex, and bariatric surgery. GEE included 54 serum samples with 27 from pre-surgery and 27 from post-surgery. mGFR Beta represents association between serum metabolite levels and mGFR. Pre-Post Beta represents change in serum metabolite level from pre-surgery to post-surgery. Serum metabolites included meet Bonferroni adjusted mGFR p-values of 4.85 × 10^−5^.

### Bariatric surgery induced serum metabolite changes

Out of 1103 metabolites, 223 (20%) were significantly increased or decreased following bariatric surgery at a Bonferroni-adjusted p-value (p=4.85×10^−5^) independent of mGFR (**Figure 1**). Of the 223 metabolites that significantly changed following bariatric surgery, 202 metabolites significantly decreased post-surgery (**Figure 1**); 227 additional metabolites were significantly associated with a change from pre-to-post bariatric surgery at an FDR threshold of 0.05.

**Figure 1.**
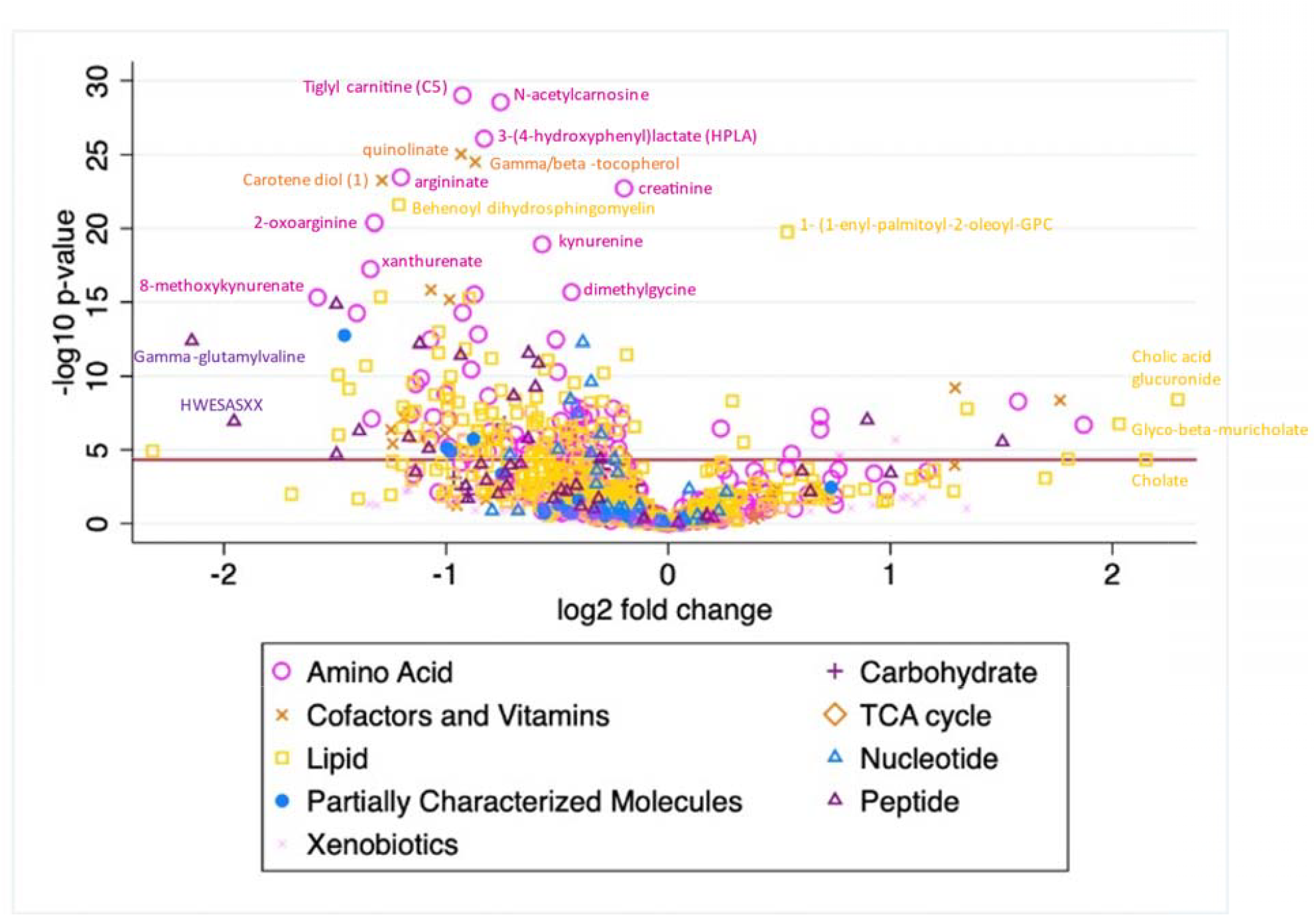
Volcano plot of mGFR-adjusted metabolite changes pre-to post-bariatric surgery analyzed by GEE adjusted for age, sex, and mGFR. X-axis demonstrates the log2(foldchange) in metabolite level from pre-surgery to post-surgery (beta-value) and Y-axis represents −log(p-value) of each metabolite. The red line signifies threshold for significant associations with bariatric surgery at a Bonferroni-adjusted p-value of 4.85 × 10^−5^.

Of the 15 most significantly altered individual metabolites (by Bonferroni-adjusted p-value), most demonstrated a significant decrease following surgery (**Table 3**). Among these, the BCAA metabolite, tiglyl carnitine (beta: −0.926, p-value: 9.89E-30) demonstrated the most statistically-significant decrease following surgery, followed by the histidine metabolite N-acetylcarnosine (beta: −0.755, p-value 2.89E-29), the tyrosine metabolite 3-(4hydroxphenyl)lactate (beta: −0.828, p-value 7.99E-27), and the nicotinate/nicotinamide metabolite quinolinate (beta: −0.935, p-value 8.22E-26). The plasmalogen metabolite, 1-(1-enyl-palmitoyl)-2-oleoyl-GPC (beta: 0.534, p-value: 1.56E-20), was associated with the most statistically significant increase following bariatric surgery. The purine metabolism metabolite, adenine, demonstrated a significant decrease following surgery (beta: −0.435, p-value: 5.05E-9). Of note, previously described uremic metabolites cresol sulfate (beta: 0.770, p-value: 2.61E-5) and TMAO (beta: 0.467, p-value: 0.0102) trended towards an increase following surgery, although were not significant per Bonferonni adjusted p-value.

**Table 3.** Serum metabolites associated with most significant changes following bariatric surgery adjusted for age, sex, and mGFR. Serum metabolite levels from the 27 post-surgery serum samples were compared with the serum metabolite levels from the 27 pre-surgery serum samples using Generalized Estimating Equations to measure magnitude of change from pre-surgery to post-surgery. Change in serum metabolite level from pre-surgery to post-surgery is indicated by the beta-value.

| Metabolite | Superpathway | Subpathway | Beta | SE | P-value | FDR |
| --- | --- | --- | --- | --- | --- | --- |
| Tiglyl carnitine (C5) | Amino Acid | Leucine, Isoleucine, Valine | -0.926 | 0.0818 | 9.89E-30 | 1.03E-26 |
| N-acetylcarnosine | Amino Acid | Histidine | -0.755 | 0.0672 | 2.89E-29 | 2.99E-26 |
| 3-(4hydroxyphenyl)lactate (HPLA) | Amino Acid | Tyrosine | -0.828 | 0.0772 | 7.99E-27 | 8.28E-24 |
| Quinolate | Cofactors and Vitamins | Nicotinate and Nicotinamide | -0.935 | 0.0890 | 8.22E-26 | 8.51E-23 |
| Gamma-tocopherol/beta-tocopherol | Cofactors and Vitamins | Tocopherol | -0.871 | 0.0838 | 2.85E-25 | 2.94E-22 |
| Argininate | Amino Acid | Urea cycle; Arginine and Proline | -1.20 | 0.119 | 3.36E-24 | 3.47E-21 |
| Carotene diol (1) | Cofactors and Vitamins | Vitamin A | -1.29 | 0.128 | 5.07E-24 | 5.24E-21 |
| Creatinine | Amino Acid | Creatine | -0.199 | 0.0199 | 1.92E-23 | 1.98E-20 |
| Behenoyl dihydrosphingomyelin | Lipid | Dihydrosphingomyelin | -1.21 | 0.125 | 2.30E-22 | 2.37E-19 |
| 2-oxoarginine | Amino Acid | Urea cycle; Arginine and Proline | -1.32 | 0.140 | 4.16E-21 | 4.28E-18 |
| 1-(1-enyl-palmitoyl)-2-oleoyl-GPC | Lipid | Plasmalogen | 0.534 | 0.0575 | 1.56E-20 | 1.61E-17 |
| Kynurenine | Amino Acid | Tryptophan | -0.568 | 0.0626 | 1.16E-19 | 1.19E-16 |
| Xanthurenate | Amino Acid | Tryptophan | -1.34 | 0.155 | 5.67E-18 | 5.81E-15 |
| Carotene diol (3) | Cofactors and Vitamins | Vitamin A | -1.07 | 0.129 | 1.32E-16 | 1.35E-13 |
| Dimethylglycine | Amino Acid | Glycine, Threonine, and Threonine | -0.435 | 0.0530 | 2.09E-16 | 2.14E-13 |

### Metabolic pathway analysis

MetaboAnalyst pathway analysis yielded four metabolite pathways demonstrating a significant upregulation and four metabolite pathways demonstrating a significant downregulation following bariatric surgery (raw p-value <0.05) (**Figure 2A and B**, respectively). Among metabolite pathways up-regulated post-bariatric surgery, the phenylacetate metabolism pathway (raw p-value 5.32E-3), followed by the bile acid biosynthesis pathway (raw p-value 7.40E-3), taurine and hypo-taurine metabolism pathway (raw p-value 9.54E-3), and porphyrin metabolism pathway (raw p-value 1.28E-2) were significantly altered. Among metabolic pathways demonstrating a significant downtrend following bariatric surgery, the alpha-linoleic acid and linoleic acid metabolism pathway (raw p-value 3.49E-4) demonstrated the most significant downtrend from pre-surgery to post-surgery. The methionine metabolism pathway (raw p-value 1.3E-2), glycine and serine metabolism pathway (raw p-value 2.66E-2), tryptophan metabolism pathway (p-value 2.66E-2), and glucose-alanine metabolism pathway (raw p-value 4.31E-2) also demonstrated significant downtrends post-surgery. Of note, only the alpha-linoleic and linoleic acid metabolism pathway demonstrated a significant downregulation per Holm p-value (3.42E-2) and FDR p-value (3.42E-2).

**Figure 2.**
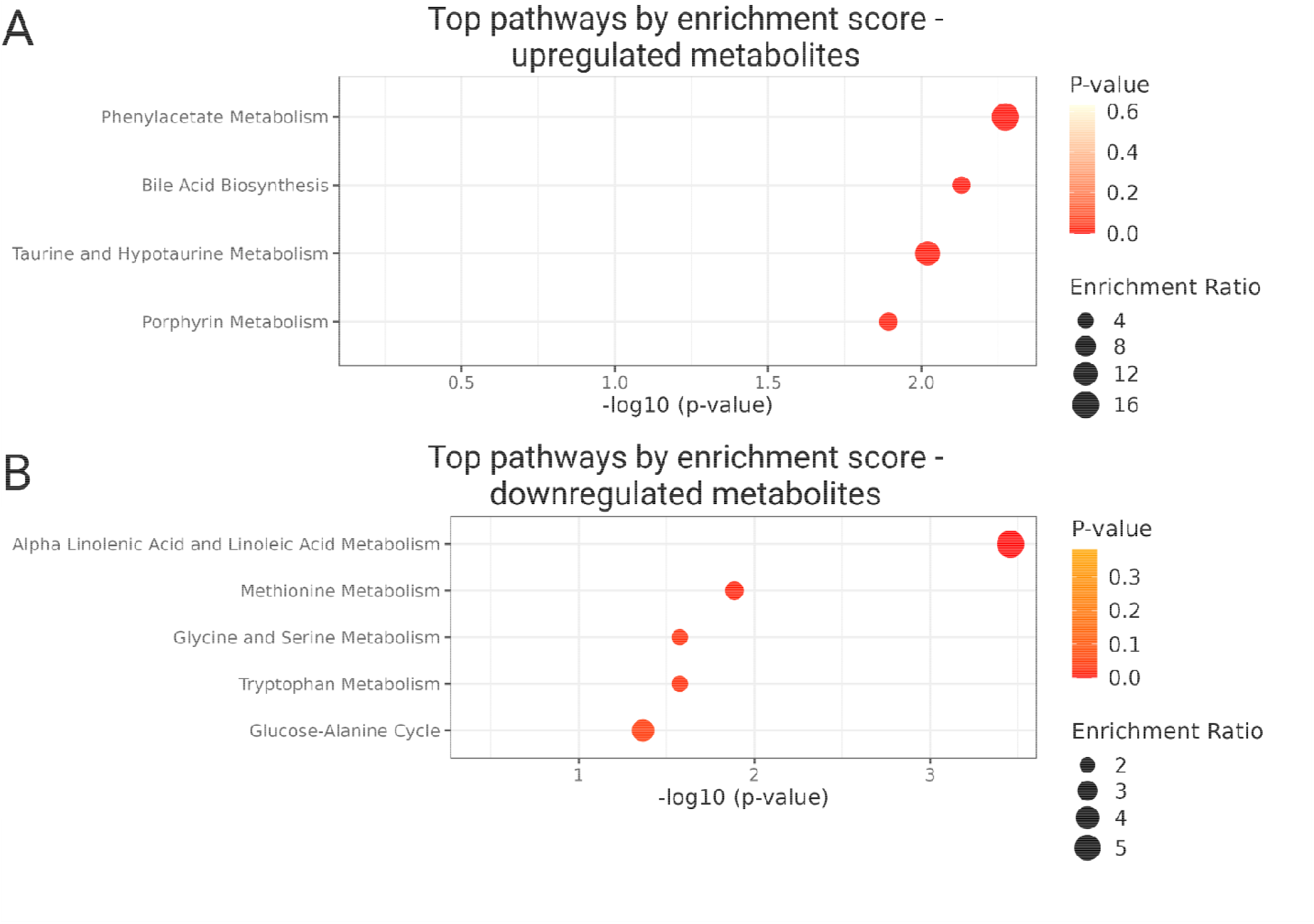
Summary plot of Over-Representation Analysis (ORA) of metabolite pathways upregulated (A) and down-regulated (B) from pre-to-post bariatric surgery. The raw p-value is represented on x-axis as −log10 (p-value) and degree of enrichment of the metabolic pathway is represented by circle size.

### Tryptophan-Kynurenine Pathway Metabolite Changes

Many metabolites involved in the inflammation-associated tryptophan-kynurenine pathway demonstrated significant decreases following bariatric surgery (**Figure 3**). Following surgery, tryptophan levels decreased (pre-post beta: −0.312, p-value: 1.75E-5). Additionally, downstream metabolites of tryptophan associated with the kynurenine catabolism pathway, such as kynurenine, 8-methoxykynurenate, N-acetylkynurenine, xanthurenic acid, and quinolinate demonstrated significant decreases post-surgery. Serotonin, a product of the non-kynurenine pathway of tryptophan metabolism, demonstrated a borderline-significant increase post-surgery (pre-post beta: 0.459, p-value: 0.034). Of note, quinolinate was the only kynurenine-pathway metabolite demonstrating a significant association with mGFR (mGFR beta: −0.702, p-value: 0.007).

**Figure 3.**
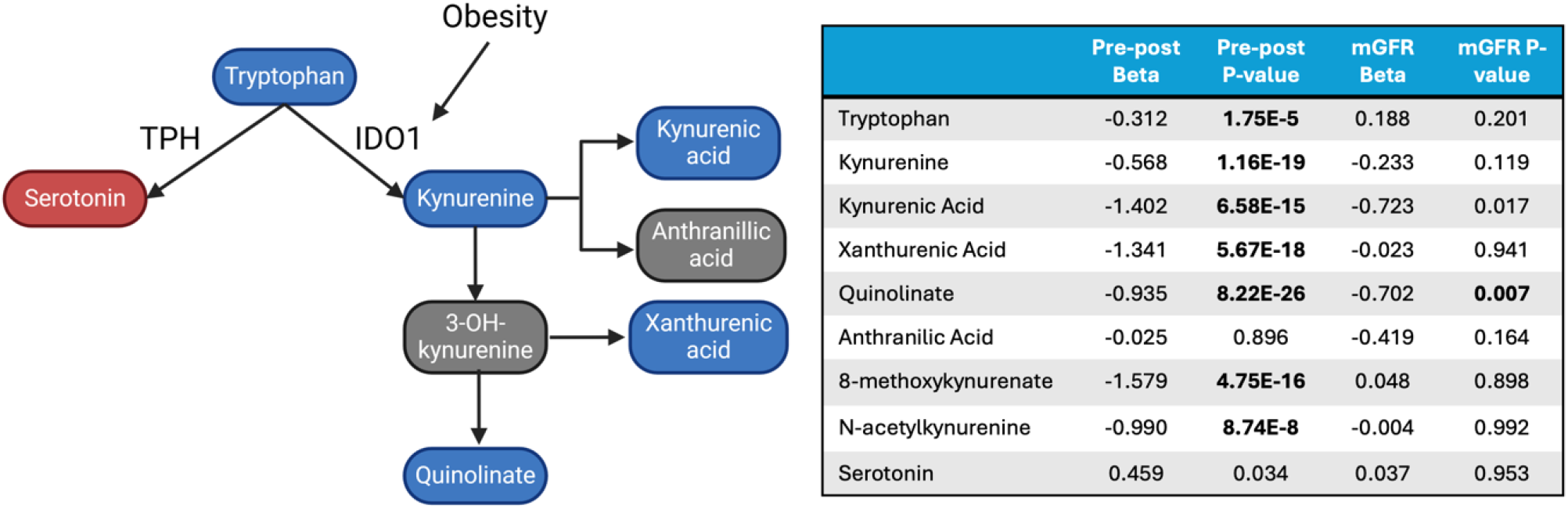
Tryptophan-Kynurenine Pathway Metabolites association with bariatric surgery and mGFR when adjusted for age, sex, and mGFR. Schema depicting catabolic pathway of tryptophan through the inflammation-associated kynurenine pathway. The accompanying table shows changes in tryptophan-kynurenine pathway metabolites from pre-to-post surgery (Pre-Post Beta) with associated p-values and metabolite associations with mGFR (mGFR Beta) and associated p-values.

### Sensitivity Analyses

Additional GEE analyses adjusting for diabetes and BMI show similar trends in serum metabolite changes following bariatric surgery (**Figure 4**). With adjustment for diabetes, 190 metabolites demonstrated a significant change following bariatric surgery per Bonferroni-adjusted p-value of 4.85E-5 and 191 metabolites demonstrated a significant post-surgery change per FDR p-value of 0.05. Similar to when unadjusted for diabetes, the most significant metabolite changes post-bariatric surgery when adjusted for diabetes included included tiglyl carnitine (beta: −0.93, p-value: 1.01E-24), argininate (beta: −1.32, p-value: 2.17E-24), N-acetylcarnosine (beta: −0.71, p-value: 3.01E-23), 3-HPLA (beta: −0.85, p-value: 1.11E-22), and quinolinate (beta: −0.95, p-value: 7.36E-21) (**Supplemental Table 1**). When adjusted for BMI, 57 metabolites demonstrated a significant change post-bariatric surgery per Bonferroni p-value of 4.85E-5 and 56 metabolites demonstrated a significant post-surgery change per FDR p-value of 0.05. The most significant surgery-related changes occurred among carotene diol (3) (beta: −1.63, p-value: 2.27E-14), carotene diol (2) (beta: −1.49, p-value: 4.86E-14), carotene diol (1) (beta: −1.65, p-value: 6.18E-14), tricosanoyl sphingomyelin (beta: −1.25, p-value: 3.92E-11), behenoyl dihydrosphingomyelin (beta: −1.31, p-value: 1.69E-9), and xanthurenate (beta: −1.38, p-value: 4.74E-9) (**Supplemental Table 2**).

**Figure 4.**
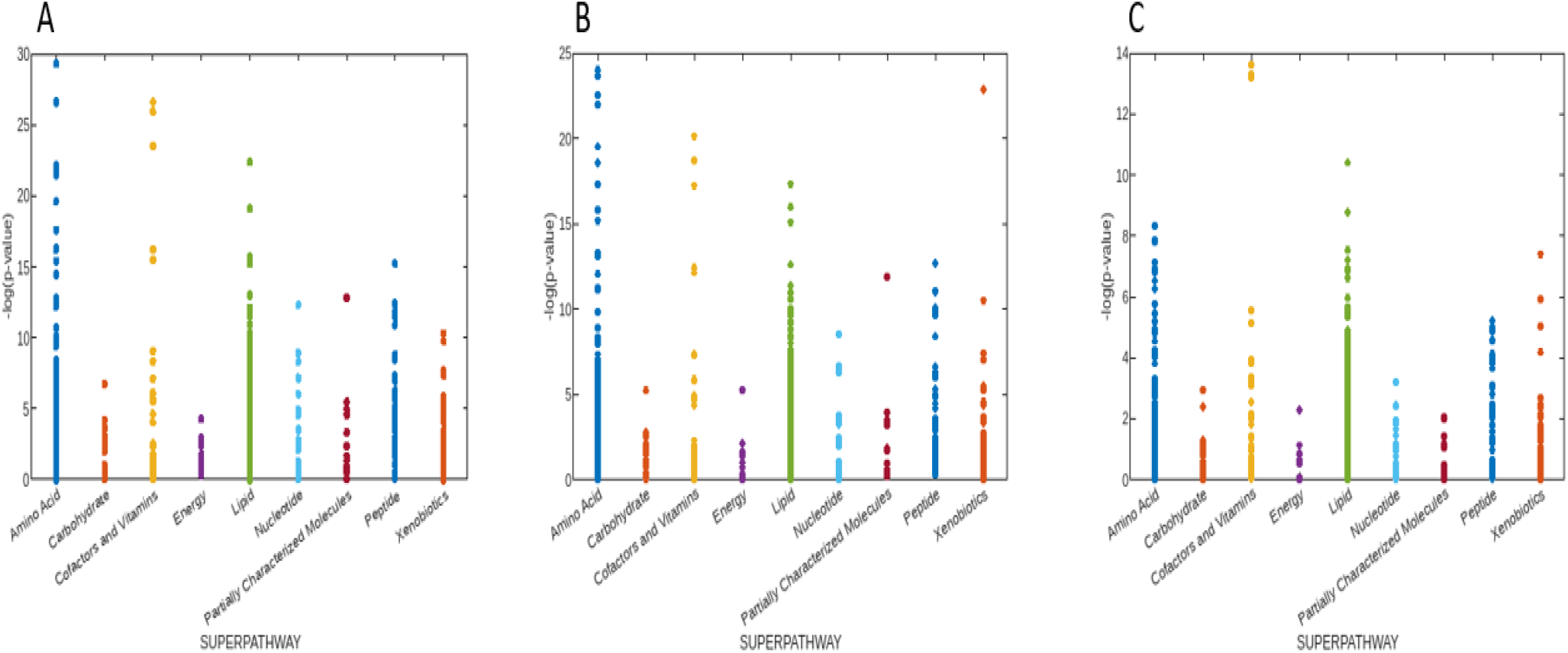
Manhattan Plots of Metabolite Changes following surgery. Data from GEE models including the terms: (A) age, sex, and mGFR; (B) age, sex, mGFR, and DM; (C) age, sex, mGFR, and BMI. Y-axis shows −log(p-value) of each metabolite and x-axis groups metabolites by metabolic superpathway.

When adjusted additionally for diabetes, TMAP (beta: −0.76, p-value: 1.17E-12) remained the most significant metabolite associated with mGFR ahead of creatinine (beta: −0.32, p-value: 1.31E-10) (**Supplemental Table 3**). Following adjustment for BMI, N-acetylthreonine, TMAP, hydroxyasparagine, and N-acetylserine all demonstrated more significant associations with mGFR than creatinine (**Supplemental Table 4**).

## DISCUSSION

Our metabolomics study investigating mGFR-adjusted metabolite changes following bariatric surgery demonstrates significant changes in specific metabolite pathways warranting further investigation. As compared with prior work in this area, this study accounts for surgery associated alterations in kidney function using adjustment by mGFR before and after bariatric surgery. Consistent with prior literature, serum metabolites associated with inflammation, pro-oxidation, and insulin sensitivity demonstrated significant decreases following bariatric surgery, even when adjusted for surgery-related mGFR changes. (1, 3) Additionally, serum metabolites associated with metabolic flexibility and anti-oxidation demonstrated significant mGFR-adjusted increases following bariatric surgery. These general trends in metabolite pathways are consistent with metabolomic changes observed in prior studies.(1, 3)

Serum levels of insulin-resistance associated metabolites, such as methionine, BCAA, histidine, tryptophan, and tyrosine, decreased following bariatric surgery when adjusted for mGFR.(1, 3, 5) Additionally, inflammation-associated metabolites, such as long-chain fatty acids, sphingomyelins, and ceramides similarly demonstrated expected decreased serum levels following bariatric surgery independent of surgery-related changes in mGFR.(1, 3) Serum metabolites associated with insulin sensitivity, such as primary bile acid metabolites, increased following surgery.(1, 3) Additionally, we noted down-regulations in the linoleic acid and carbohydrate-alanine pathways, which correlate to the malabsorptive properties and decreases in caloric intake associated with bariatric surgery.(1)

The significant decrease in serum kynurenine-pathway-related metabolites demonstrates one serum metabolite pathway change that may contribute to improved kidney outcomes following surgery. Serum metabolite levels of kynurenine pathway metabolites have been previously associated with kidney disease, chronic inflammation, malignancy, and cardiovascular disease.(5, 7, 8, 19) Elevated kynurenine pathway metabolites have also been associated with development of AKI and CKD progression.(7, 8, 19) In our study, the serum levels of kynurenine metabolites, including, kynurenine, xanthurenate, 8-methoxykynurenate, kynurenate, N-acetylkynurenine, and quinolinate, significantly decreased following bariatric surgery. Despite the decrease in kynurenine-pathway metabolites post-surgery, our study did not show evidence of an association between these serum metabolites and mGFR, suggesting that at least some metabolic effects are related to surgery rather than changes in kidney function.

Our study demonstrated a significant mGFR-independent decrease in adenine levels following bariatric surgery. This is a second metabolite pathway that may connect obesity to CKD progression. Adenine has been implicated in the progression of diabetic kidney disease. (20) Animal models suggest that exogenous adenine administration results in kidney damage through a variety of mechanisms, including glomerulosclerosis, interstitial fibrosis, tubular atrophy, and inflammation. (21, 22, 23)

Our study also implicates the taurine and hypotaurine pathway, which was upregulated post-surgery in our pathway analysis, as a possible contributor to the improved kidney function outcomes associated with bariatric surgery. Taurine is an essential amino acid inversely proportional to abdominal obesity, waist-to-hip ratio, and BMI. (24) Further, studies implicate taurine in ameliorating obesity by stimulating energy expenditure, modulating metabolism of lipids, and providing anti-inflammatory and anti-oxidant properties. (25) Numerous studies suggest that taurine also plays a role in regulating renal blood flow, osmoregulation, preventing ischemia and reperfusion injury, and antioxidation. (26) Furthermore, individuals with CKD and diabetic nephropathy have lower levels of serum taurine than individuals with normal kidney function, suggesting a protective function of taurine against kidney disease. (26, 27)

We observed significant increases among serum levels of the minimally studied plasmalogen metabolites. Serum plasmalogen metabolites, 1-(1-enyl-palmitoyl)-2-oleoyl-GPC and 1-(1-enyl-palmitoyl)-2-palmitoleoyl-GPC, demonstrated significant increases following bariatric surgery. Although not previously studied in the context of kidney function, plasmalogens have been implicated in ameliorating the progression of several degenerative and metabolic disorders, including Alzheimer’s disease, obesity, and coronary artery disease, given their anti-oxidant and anti-inflammatory properties. (28, 29)

One factor associated with several of the metabolic changes noted in this study may include the marked shift in gut microbiome caused by bariatric surgery. (3) Several studies suggest that bariatric surgery leads to gut microbial changes that resemble a “lean microbiome” phenotype compared to a pre-surgery “obese microbiome” phenotype. (3) The increase among many of the anti-oxidant lipid plasmalogen metabolites and bile acid biosynthesis metabolite following bariatric surgery may be associated with surgery-related gut microbiome alterations, as these metabolites are largely synthesized by a subset of gut bacteria. (3, 30) However, the same microbiome alterations in bariatric surgery patients may also explain the paradoxical increase in the pro-inflammatory gut-derived metabolites TMAO and p-cresol sulfate. (31, 32) We also observed significant increases in the serum phenylacetate-derived metabolites, such as the cardiac disease-associated metabolite phenylacetylglutamine (PAG), which have previously been associated with gut microbiome changes. (33, 34) These metabolite changes suggest a possible association to the altered gastrointestinal anatomy and gut microbiome resulting from bariatric surgery.

This study identified the serum metabolite, N, N, N-trimethyl-alanylproline betaine (TMAP) as most significantly associated with mGFR. Strikingly, this association was stronger than that of creatinine with mGFR. TMAP has previously been demonstrated to be a marker of kidney function in patients with early CKD, ESRD, and requiring hemodialysis. (35) Additionally, our study confirms the results of prior studies suggesting associations between mGFR and the serum metabolites pseudouridine, N-acetylserine, myo-inositol, and erythronate, although 3 of these metabolites (N-acetylthreonine, N-acetylserine, myo-inositol) significantly decrease post-bariatric surgery, which may limit their roles as filtration markers in bariatric surgery patients. (10, 36–38) Further studies of these serum metabolites in other bariatric surgery cohorts will be helpful to evaluate their role in estimating kidney function in this setting.

There are several limitations to the study, most notably the small sample size of 27 patients and limited post-surgery follow-up duration, which is likely why some known metabolites, such as serum ketone bodies and short-chain fatty acids, did not reach Bonferroni-corrected significance. Additionally, the sample was composed of a largely homogenous population of white patients. Further, post-surgery serum sample collections were at 6 months, limiting our ability to understand whether identified serum metabolites continue to be significantly decreased or increased beyond the time points available in the study, and long-term relationships between these metabolites and hard end-organ outcomes. Finally, because our results demonstrate relative changes in metabolite levels, rather than absolute changes, metabolites with very low pre-surgery levels with minimal changes post-surgery may yield “significant changes” on analysis, producing false positive results. (39)

## CONCLUSION

This study demonstrates mGFR-independent changes in several serum metabolite pathways that may explain the salutary effects of bariatric surgery on kidney function. In general, serum metabolites associated with inflammation, pro-oxidation, and insulin resistance decrease following surgery, whereas serum metabolites associated with anti-oxidation, anti-inflammation, and improved glucose utilization increase following surgery. These findings are consistent with the results of prior work in this area, while adjusting for changes in kidney function that may occur following bariatric surgery. Our study re-emphasizes the potential relationship of kynurenine, adenine, and taurine/hypotaurine metabolites with kidney function and identifies novel metabolites, such as pro-oxidative plasmalogens, associated with bariatric surgery. Further studies investigating the mechanistic role of kynurenine metabolites, adenine, taurine/hypotaurine metabolites, and plasmalogen metabolites may clarify the mechanisms of bariatric surgery-induced improved kidney outcomes. Additionally, further study of the metabolite TMAP may identify its use as an early correlate of kidney function among patients undergoing bariatric surgery.

## Data Availability

All data produced in the present study are available upon reasonable request to the authors and subject to a data user agreement.

## ACKNOWLEDGEMENTS

This research was supported by the National Institutes of Health (National Institute of Diabetes, Digestive, and Kidney Diseases) grants K23 DK108515-01 and R01DK097020.

