## Supplemental Materials for "Serum Metabolomic Changes Following Bariatric Surgery Adjusted for Measured Glomerular Filtration Rate Suggest Mechanisms of Kidney Protection"

Supplemental Tables and Figures:

**Supplemental Table 1. Single serum metabolites associated with most significant changes following bariatric surgery adjusted for age, sex, mGFR, and diabetes mellitus.** Serum metabolite levels from the 27 post-surgery serum samples were compared with the serum metabolite levels from the 27 pre-surgery serum samples using Generalized Estimating Equations with adjustments for age, sex, mGFR, and diabetes mellitus to measure magnitude of change from pre-surgery to post-surgery. Change in serum metabolite level from pre-surgery to post-surgery is indicated by the beta-value.

| **Metabolite** | **Superpathway** | **Subpathway** | **Beta** | **SE** | **P-value** | **FDR** |
| --- | --- | --- | --- | --- | --- | --- |
| Tiglyl carnitine (C5) | Amino Acid | Leucine, Isoleucine, Valine | -0.93 | 0.09 | 1.01E-24 | 1.05E-21 |
| Argininate | Amino Acid | Urea cycle; Arginine and Proline | -1.32 | 0.13 | 2.17E-24 | 2.24E-21 |
| N-acetylcarnosine | Amino Acid | Histidine | -0.71 | 0.07 | 3.01E-23 | 3.10E-20 |
| 3-(4hydroxphenyl)lactate (HPLA) | Amino Acid | Tyrosine | -0.85 | 0.08 | 1.11E-22 | 1.15E-19 |
| Quinolinate | Cofactors | Nicotinate and Nicotinamide | -0.95 | 0.10 | 7.36E-21 | 7.59E-18 |
| Creatinine | Amino Acid | Creatine | -0.17 | 0.02 | 3.11E-20 | 3.20E-17 |
| Carotene diol (1) | Cofactors and Vitamins | Vitamin A | -1.28 | 0.14 | 2.06E-19 | 2.12E-16 |
| 2-oxoarginine | Amino Acid | Urea cycle; Arginine and Proline | -1.42 | 0.16 | 2.78E-19 | 2.86E-16 |
| Behenoyl dihdrosphingomyelin | Lipid | Dihydrosphingomyelins | -1.21 | 0.14 | 4.68E-18 | 4.80E-15 |
| Kynurenine | Amino Acid | Tryptophan | -0.61 | 0.07 | 4.92E-19 | 5.04E-15 |
| Gamma-tocopherol/beta-tocopherol | Cofactors | Tocopherol | -0.82 | 0.10 | 5.86E-18 | 6.00E-15 |
| 2S,3R-dihydroxybutrate | Lipid | Fatty Acid, Dihydroxy | -1.44 | 0.17 | 1.08E-16 | 1.11E-13 |
| Cystathionine | Amino Acid | Methionine, Cysteine, SAM and Taurine | -1.22 | 0.15 | 1.63E-16 | 1.66E-13 |
| Xanthurenate | Amino Acid | Tryptophan | -1.40 | 0.17 | 6.67E-16 | 6.81E-13 |
| 1-(1-enyl-palmitoyl)-2-oleoyl-GPC | Lipid | Plasmalogen | 0.52 | 0.07 | 8.15-16 | 8.31E-13 |

**Supplemental Table 2. Single serum metabolites associated with most significant changes following bariatric surgery adjusted for age, sex, mGFR, and BMI.** Serum metabolite levels from the 27 post-surgery serum samples were compared with the serum metabolite levels from the 27 pre-surgery serum samples using Generalized Estimating Equations with adjustments for age, sex, mGFR, and BMI to measure magnitude of change from pre-surgery to post-surgery. Change in serum metabolite level from pre-surgery to post-surgery is indicated by the beta-value.

| **Metabolite** | **Superpathway** | **Subpathway** | **Beta** | **SE** | **P-value** | **FDR** |
| --- | --- | --- | --- | --- | --- | --- |
| Carotene diol (1) | Cofactors and Vitamins | Vitamin A | -1.63 | 0.21 | 2.27E-14 | 2.35E-11 |
| Carotene diol (2) | Cofactors and Vitamins | Vitamin A | -1.49 | 0.19 | 4.86E-14 | 5.02E-11 |
| Carotene diol (3) | Cofactors and Vitamins | Vitamin A | -1.65 | 0.22 | 6.18E-14 | 6.38E-11 |
| Tricosanoyl sphingomyelin | Lipid | Sphingomyelins | -1.25 | 0.18 | 3.92E-11 | 4.04E-8 |
| Behenoyl dihydrosphingomyelin | Liipd | Dihydrosphingomyelins | -1.31 | 0.21 | 1.69E-9 | 1.74E-06 |
| Xanthurenate | Amino Acid | Tryptophan | -1.38 | 0.23 | 4.74E-9 | 4.88E-6 |
| 8-methoxykynurenate | Amino Acid | Tryptophan | -1.66 | 0.29 | 1.37E-8 | 1.41E-5 |
| 2-oxoarginine | Amino Acid | Urea Cycle; Arginine and Proline | -1.30 | 0.23 | 1.55E-8 | 1.59E-5 |
| Lignoceroyl sphingomyelin | Lipid | Sphingomyelins | -0.95 | 0.17 | 2.98E-8 | 3.06E-5 |
| sphingomyelin | Lipid | Sphingomyelins | -0.99 | 0.18 | 6.36E-8 | 6.51E-5 |
| tiglyl carnitine (C5) | Amino Acid | Leucine, Isoleucine, Valine | -0.710 | 0.13 | 7.55E-8 | 7.72E-5 |
| prolylhydroxyproline | Amino Acid | Urea Cycle; Arginine and Proline | 1.01 | 0.19 | 1.15E-7 | 1.17E-4 |
| 2S,3R-dihydroxybutrate | Lipid | Fatty Acid, Dihydroxy | -1.09 | 0.20 | 1.22E-7 | 1.25E-4 |
| Behenoyl sphingomyelin | Lipid | Sphingomyelins | -0.71 | 0.13 | 1.37E-7 | 1.40E-4 |
| Alpha-ketobutyrate | Amino Acid | Methionine, Cysteine, SAM and Taurine | -1.57 | 0.30 | 1.56E-7 | 1.59E-4 |

**Supplemental Table 3. Top 15 serum metabolites significantly associated with mGFR adjusted for age, sex, and diabetes mellitus.** GEE included 54 serum samples with 27 from pre-surgery and 27 from post-surgery with adjustments for age, sex, and diabetes mellitus. Beta value represents the association between serum metabolite levels and mGFR.

| **Metabolite** | **Subpathway** | **Beta** | **SE** | **p-value** |
| --- | --- | --- | --- | --- |
| N,N,N-trimethyl-alanylproline betaine (TMAP) | Amino Acid | -0.763 | 0.107 | 1.17E-12 |
| creatinine | Amino Acid | -0.320 | 0.050 | 1.31E-10 |
| erythronate | Carbohydrate | -0.617 | 0.098 | 3.54E-10 |
| N-acetyl threonine | Amino Acid | -0.760 | 0.138 | 3.84E-8 |
| 5,6-dihydrouridine | Nucleotide | -0.827 | 0.152 | 4.93E-8 |
| pseudouridine | Nucleotide | -0.877 | 0.163 | 7.30E-8 |
| 5-methylthioribose | Amino Acid | -0.531 | 0.099 | 8.09E-8 |
| Myo-inositol | Lipid | -0.659 | 0.128 | 2.42E-7 |
| N-acetylserine | Amino Acid | -0.494 | 0.097 | 3.36E-7 |
| N6-succinyladenosine | Nucleotide | -1.252 | 0.246 | 3.57E-7 |
| N-acetylneuraminate | Carbohydrate | -0.557 | 0.113 | 8.72E-7 |
| pyroglutamine | Amino Acid | -1.666 | 0.351 | 2.24E-6 |
| C-glycosyltryptophan | Amino Acid | -0.619 | 0.132 | 2.98E-6 |
| N1-methylinosine | Amino Acid | -0.904 | 0.194 | 3.36E-6 |
| Glu-gly-asn-val | Peptide | -0.943 | 0.207 | 5.25E-6 |

**Supplemental Table 4. Top 15 serum metabolites significantly associated with mGFR adjusted for age, sex, and body mass index (BMI).** GEE included 54 serum samples with 27 from pre-surgery and 27 from post-surgery with adjustments for age, sex, and BMI. Beta value represents the association between serum metabolite levels and mGFR.

| **Metabolite** | **Subpathway** | **Beta** | **SE** | **p-value** |
| --- | --- | --- | --- | --- |
| N-acetyl threonine | Amino Acid | -0.840 | 0.111 | 4.77E-14 |
| N,N,N-trimethyl-alanylproline betaine (TMAP) | Amino Acid | -0.742 | 0.102 | 4.57E-13 |
| Hydroxyasparagine | Amino Acid | -0.775 | 0.114 | 1.23E-11 |
| N-acetylserine | Amino Acid | -0.541 | 0.085 | 2.41E-10 |
| creatinine | Amino Acid | -0.353 | 0.061 | 3.46E-10 |
| pseudouridine | Nucleotide | -0.899 | 0.151 | 2.76E-9 |
| Myo-inositol | Lipid | -0.692 | 0.118 | 3.90E-9 |
| N1-methylinosine | Nucleotide | -0.989 | 0.181 | 4.87E-8 |
| N6-succinyladenosine | Nucleotide | -1.263 | 0.232 | 5.19E-8 |
| 5,6-dihydrouridine | Nucleotide | -0.815 | 0.151 | 6.19E-8 |
| erythronate | Carbohydrate | -0.590 | 0.111 | 1.06E-7 |
| N-acetylneuraminate | Carbohydrate | -0.566 | 0.109 | 1.99E-7 |
| N-carbomoylvaline | Amino Acid | -0.949 | 0.187 | 3.68E-7 |
| 5-methylthioribose | Amino Acid | -0.563 | 0.111 | 3.97E-7 |
| O-sulfo-L-tyrosine | Amino Acid | -0.628 | 0.129 | 1.09E-6 |
